# Effectiveness and Duration of Protection of a Fourth Dose of COVID-19 mRNA Vaccine among Long-Term Care Residents in Ontario, Canada

**DOI:** 10.1101/2022.09.29.22280526

**Authors:** Ramandip Grewal, Lena Nguyen, Sarah A Buchan, Sarah E Wilson, Andrew P Costa, Jeffrey C Kwong

**Author notes:** **Correspondence to:** Jeffrey C Kwong, Senior Scientist, ICES, G1 06, 2075 Bayview Avenue, Toronto, Ontario, Canada, M4N 3M5, (or @DrJeffKwong on Twitter.

## Abstract

**Background:** As of December 30, 2021, Ontario long-term care (LTC) residents who received a third dose of COVID-19 vaccine ≥84 days previously were offered a fourth dose to prevent a surge in COVID-19-related morbidity and mortality due to the Omicron variant. Seven months have passed since fourth doses were implemented, allowing for the examination of fourth dose protection over time.

**Methods:** We used a test-negative design and linked databases to estimate the marginal effectiveness (4 versus 3 doses) and vaccine effectiveness (VE; 2, 3, or 4 doses versus no doses) of mRNA vaccines among Ontario LTC residents aged ≥60 years who were tested for SARS-CoV-2 between December 30, 2021 and August 3, 2022. Outcome measures included any Omicron infection, symptomatic infection, and severe outcomes (hospitalization or death).

**Results:** We included 21,275 Omicron cases and 273,466 test-negative controls. The marginal effectiveness of a fourth dose <84 days ago compared to a third dose received ≥84 days ago was 23% (95% Confidence Interval [CI] 17-29%), 36% (95%CI 26-44%), and 37% (95%CI 24-48%) against SARS-CoV-2 infection, symptomatic infection, and severe outcomes, respectively. Additional protection provided by a fourth dose compared to a third dose was negligible against all outcomes ≥168 days after vaccination. Compared to unvaccinated individuals, vaccine effectiveness (VE) of a fourth dose decreased from 49% (95%CI 44%-54%) to 18% (95%CI 5-28%) against infection, 69% (95%CI 62-75%) to 44% (95%CI 24-59%) against symptomatic infection, and 82% (95%CI 77-86%) to 74% (95%CI 62-82%) against severe outcomes <84 days versus ≥168 days after vaccination.

**Conclusions:** Our findings suggest that fourth doses of mRNA COVID-19 vaccines provide additional protection against Omicron-related outcomes in LTC residents, but the protection wanes over time, with more waning seen against infection than severe outcomes.

## INTRODUCTION

On December 30, 2021, Ontario, Canada began offering fourth (second booster) doses to long-term care residents (LTC) who had received their third dose at least 3 months (≥84 days) prior.^1^ The preferred product was 100 micrograms (mcg) of mRNA-1273 (Moderna Spikevax), an off-label recommendation for this dosage as a booster dose.^1^ Previous analyses on vaccine effectiveness (VE) compared to unvaccinated individuals and marginal effectiveness of fourth versus third doses conducted among Ontario LTC residents found that a fourth dose received ≥7 days ago increased protection against Omicron-related SARS-CoV-2 infection, symptomatic infection, and severe outcomes.^2^ With over 7 months having passed since fourth doses were implemented for this population, the objectives of this study were to estimate marginal effectiveness of fourth doses compared to third doses and VE of 2, 3, and 4 doses across different time periods since vaccination and explore potential waning of fourth dose protection.

## METHODS

We conducted a test-negative study among LTC residents aged ≥60 years as of December 30, 2021 (date eligible for fourth doses) across 626 LTC facilities in Ontario using laboratory, vaccination, reportable disease, and health administrative data from December 30, 2021 to August 3, 2022. We excluded individuals testing positive for SARS-CoV-2 ≤90 days ago, individuals who received a fourth dose before December 30, 2021, and those who ever received a non-mRNA vaccine. Since Omicron was the dominant variant of concern during our study period, cases were considered to be Omicron unless confirmed as Delta (B.1.617.2) through whole genome sequencing or S-gene target failure.^3–5^

We estimated VE (2, 3, and 4 doses versus unvaccinated) and marginal effectiveness (4 doses versus 3 doses received ≥84 days ago) against infection (SARS-CoV-2-positive individuals, irrespective of symptoms), symptomatic infection (individuals with ≥1 symptom consistent with COVID-19 disease that was recorded in the Ontario Laboratories Information System [OLIS]^2^ when tested), and severe outcomes (hospitalization or death due to, or partially due to, COVID-19). Cases and controls were sampled by week of test. If cases had multiple infections, the first infection was selected. The first negative test within a week was selected for controls. Controls could later be considered cases if testing negative in previous weeks but subsequently testing positive later in the study period. Further details on LTC testing practices in Ontario and the methods and sampling strategy have been detailed elsewhere.^2^

We used multivariable logistic regression to compare the odds of vaccination in cases to test-negative controls while adjusting for age, sex, public health region of residence, week of test, whether they had a SARS-CoV-2 test >90 days prior, and number of comorbidities. We used a generalized estimating equations framework with an exchangeable correlation structure to account for clustering at the facility level. We calculated VE and marginal effectiveness using the formula 1-adjusted odds ratio. Datasets were linked using unique encoded identifiers and analyzed at ICES (formerly the Institute of Clinical Evaluative Sciences).

### Ethics approval

ICES is a prescribed entity under Ontario’s Personal Health Information Protection Act (PHIPA). Section 45 of PHIPA authorizes ICES to collect personal health information, without consent, for the purpose of analysis or compiling statistical information with respect to the management of, evaluation or monitoring of, the allocation of resources to or planning for all or part of the health system. Projects that use data collected by ICES under section 45 of PHIPA, and use no other data, are exempt from REB review. The use of the data in this project is authorized under section 45 and approved by ICES’ Privacy and Legal Office.

## RESULTS

During the study period, 92.0% (66,994 of 72,846) of Ontario LTC residents were tested for SARS-CoV-2 at least once. At the time of testing, 46.3% and 48.7% of cases and controls, respectively, had received a fourth dose (Table S1). Fewer residents with a fourth dose resided in a facility with an active outbreak compared to residents who received a third dose ≥84 days ago (Table S2). Among adults who received at least a third dose, 57% received mRNA-1723 for the third dose. Among those with a fourth dose, 95% received mRNA-1273.

Compared to individuals who received a third dose ≥84 days ago, marginal effectiveness of a fourth dose received <84 days ago was 23% (95%CI 17-29%) against infection, 36% (95%CI 26-44%) against symptomatic infection, and 37% (95%CI 24-48%) against severe outcomes (Figure 1, Table S3). Marginal effectiveness decreased as time since fourth dose increased, with negligible (<10%) additional protection against infection and symptomatic infection by 112-139 days and against severe outcomes after 168 days.

**Figure 1:**
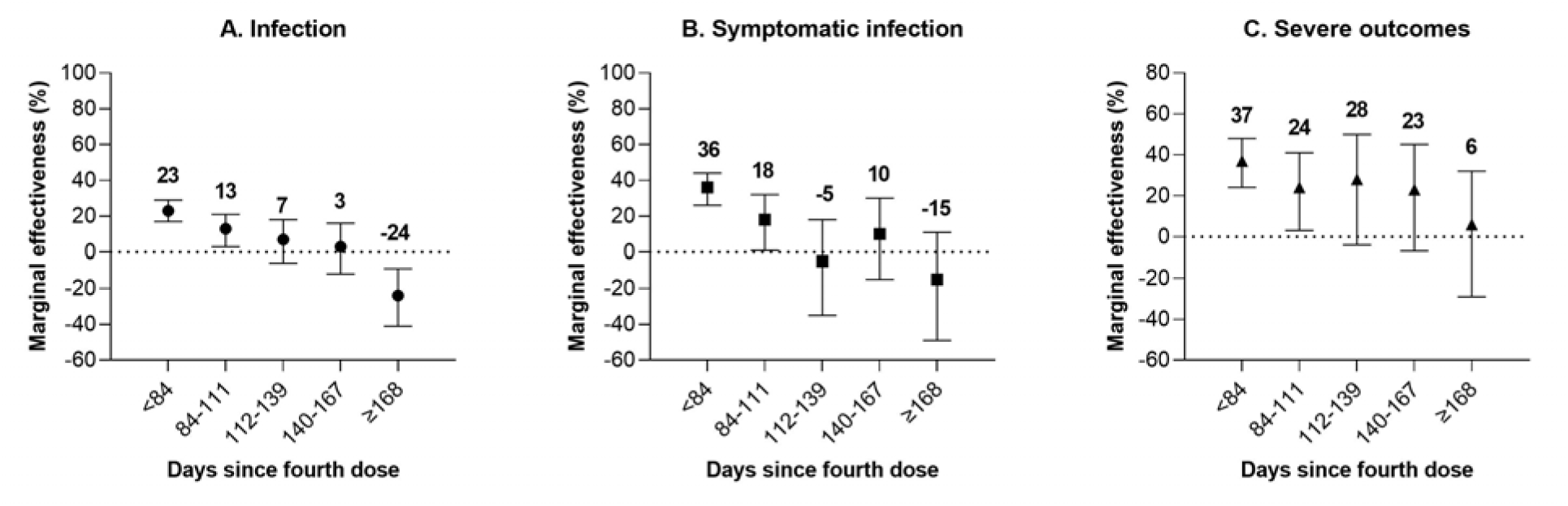
Marginal effectiveness of a fourth mRNA COVID-19 vaccine dose against Omicron outcomes compared to a third dose received ≥84 days ago among long-term care (LTC) residents in Ontario, Canada.

Compared to unvaccinated individuals, VE increased with each additional dose received but decreased as time since vaccination increased (Figure 2, Table S4). VE was highest at <84 days post-fourth dose (49% [95%CI 44-54%] against infection, 69% [95%CI 61-75%] against symptomatic infection, and 82% [95%CI 77-86%] against severe outcomes). At ≥168 days after fourth dose receipt, VE decreased to 18% (95%CI 5-28%) against infection, 44% (95%CI 24-59%) against symptomatic infection, and 74% (95%CI 62-82%) against severe outcomes.

**Figure 2:**
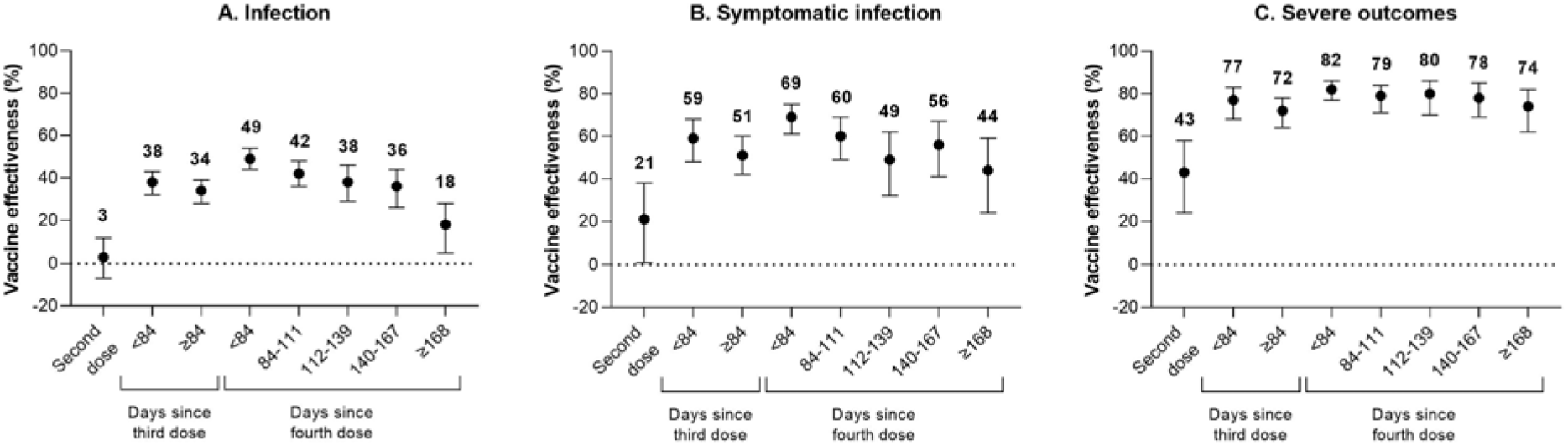
Vaccine effectiveness (VE) of a fourth mRNA COVID-19 vaccine dose against Omicron outcomes compared to unvaccinated individuals among long-term care (LTC) residents in Ontario, Canada.

## DISCUSSION

Among LTC residents in Ontario, the effectiveness of fourth doses of mRNA COVID-19 vaccines against Omicron outcomes waned over time and was highest shortly after fourth dose receipt. Nonetheless, compared to unvaccinated individuals, fourth dose VE was maintained even ≥168 days after vaccination, with VE against severe outcomes of 74%. Compared to a third dose received ≥84 days ago, we found that the boost in effectiveness received from a fourth dose (i.e., marginal effectiveness) against all outcomes was gone 6 months (168 days) after vaccination. The most rapid decline in marginal effectiveness over time was seen against SARS-CoV-2 infection, followed by symptomatic infection; there was no additional protection of a fourth dose against these outcomes after 4 months (112 days).

Similar to other studies on fourth doses,^6,7^ the degree of waning against severe outcomes was lower than what was seen against infection. Compared to unvaccinated individuals, VE ≥168 days after fourth dose receipt (74%) returned to similar levels seen ≥84 days after receiving a third dose (72%). In comparison, protection against infection and symptomatic infection reached comparable levels between 112-167 days after a fourth dose. Though direct comparisons to other studies are difficult due to differences in time periods, outcome definitions, and methodology, our findings of fourth dose waning against severe outcomes are similar to those seen among a LTC facility cohort in Sweden where marginal effectiveness of an mRNA fourth dose (compared to a third dose) against all-cause mortality was 39% 7-60 days after receiving a fourth dose, and declined to 27% 61-126 days after vaccination.^8^ In our study, we found marginal effectiveness against severe outcomes when compared to a third dose received ≥84 days ago reduced from 37% <84 days after fourth dose receipt to 24% 84-111 days after vaccination.

Limitations of this study include the potential for residual confounding, the possibility of some Delta cases being included in the analyses because not all specimens underwent S-gene target failure screening or whole genome sequencing, challenges in distinguishing between waning and increased immune invasion of new circulating Omicron subvariants, inability to determine the dosage (100 or 50 mcg) of fourth doses mRNA-1273 received, and our symptomatic cohort being limited to individuals who had symptoms recorded in OLIS. Fourth doses increased protection against Omicron outcomes compared to a third dose and compared to unvaccinated individuals, but jurisdictions must monitor time since fourth receipt among LTC residents to help subsequent booster planning to ensure residents remain protected against COVID-19 and related outcomes. To continue to provide increased protection against SARS-CoV-2 infection for this vulnerable population, an additional booster may be needed 4-6 months after fourth dose receipt. Protection against severe outcomes is more stable; residents seem to maintain decent protection even 6 months after receiving a fourth dose.

## Data Availability

The dataset from this study is held securely in coded form at ICES. While legal data sharing agreements between ICES and data providers (e.g., healthcare organizations and government) prohibit ICES from making the dataset publicly available, access may be granted to those who meet pre-specified criteria for confidential access, available at www.ices.on.ca/DAS.

## Code availability

The full dataset creation plan and underlying analytic code are available from the authors upon request, understanding that the computer programs may rely upon coding templates or macros that are unique to ICES and are therefore either inaccessible or may require modification.

## Acknowledgements

We would like to acknowledge Public Health Ontario for access to vaccination data from COVaxON, case-level data from CCM and COVID-19 laboratory data, as well as assistance with data interpretation. We also thank the staff of Ontario’s public health units who are responsible for COVID-19 case and contact management and data collection within CCM. We thank IQVIA Solutions Canada Inc. for use of their Drug Information File. The authors are grateful to the Ontario residents without whom this research would be impossible. We would also like to acknowledge Sharifa Nasreen for producing the figures for this manuscript.

## Funding and disclaimers

This work was supported by the Applied Health Research Questions (AHRQ) Portfolio at ICES, which is funded by the Ontario Ministry of Health (MOH). For more information on AHRQ and how to submit a request, please visit www.ices.on.ca/DAS/AHRQ. This work was also supported by the Ontario Health Data Platform (OHDP), a Province of Ontario initiative to support Ontario’s ongoing response to COVID-19 and its related impacts. This work was supported by Public Health Ontario. This study was also supported by ICES, which is funded by an annual grant from the Ontario MOH and the Ministry of Long-Term Care (MLTC). The study sponsors did not participate in the design and conduct of the study; collection, management, analysis and interpretation of the data; preparation, review or approval of the manuscript; or the decision to submit the manuscript for publication. Parts of this material are based on data and/or information compiled and provided by the Canadian Institute for Health Information (CIHI), and by Ontario Health (OH). However, the analyses, conclusions, opinions and statements expressed herein are solely those of the authors, and do not reflect those of the funding or data sources; no endorsement by ICES, MOH, MLTC, OHDP, its partners, the Province of Ontario, Ontario Health, CIHI, or OH is intended or should be inferred.

