## Supplementary material for "Effectiveness and Duration of Protection of a Fourth Dose of COVID-19 mRNA Vaccine among Long-Term Care Residents in Ontario, Canada"

**Supplementary Appendix**

This appendix has been provided by the authors to give readers additional information about their work.

**Table S1:** Descriptive characteristics of long-term care (LTC) residents tested for SARS-CoV-2 between December 30, 2021 and August 3, 2022 in Ontario, Canada, comparing Omicron cases to SARS-CoV-2-negative controls

|  | **SARS-CoV-2 negative, n (%)^a^** | **Omicron, n (%)^a^** | **SD^b^** |
| --- | --- | --- | --- |
| **Total** | 273,466 | 21,275 |  |
| Characteristics |  |  |  |
| Exposure |  |  |  |
| Unvaccinated | 7,005 (2.6%) | 805 (3.8%) | 0.07 |
| 1 dose received | 1,129 (0.4%) | 110 (0.5%) | 0.02 |
| 2 doses received | 12,472 (4.6%) | 1,442 (6.8%) | 0.10 |
| 3 doses received <84 days prior to test | 28,133 (10.3%) | 1,842 (8.7%) | 0.06 |
| 3 doses received ≥84 days prior to test | 91,251 (33.4%) | 7,232 (34.0%) | 0.01 |
| 4 doses received <84 days prior to test | 77,695 (28.4%) | 3,736 (17.6%) | 0.26 |
| 4 doses received 84-111 days prior to test | 21,450 (7.8%) | 1,915 (9.0%) | 0.04 |
| 4 doses received 112-139 days prior to test | 13,253 (4.8%) | 1,094 (5.1%) | 0.01 |
| 4 doses received 140-167 days prior to test | 10,223 (3.7%) | 955 (4.5%) | 0.04 |
| 4 doses received ≥168 days prior to test | 10,855 (4.0%) | 2,144 (10.1%) | 0.24 |
| Male sex | 83.39 ± 9.54 | 83.98 ± 9.32 | 0.06 |
| Age (years; mean SD^c^) | 27,892 (10.2%) | 1,872 (8.8%) | 0.05 |
| 60 to 69 | 58,677 (21.5%) | 4,342 (20.4%) | 0.03 |
| 70 to 79 | 186,897 (68.3%) | 15,061 (70.8%) | 0.05 |
| ≥80 | 87,220 (31.9%) | 7,257 (34.1%) | 0.05 |
| Public health unit region |  |  |  |
| Central East | 19,773 (7.2%) | 1,540 (7.2%) | 0.00 |
| Central West | 44,278 (16.2%) | 4,623 (21.7%) | 0.14 |
| Durham | 9,977 (3.6%) | 699 (3.3%) | 0.02 |
| Eastern | 24,982 (9.1%) | 1,785 (8.4%) | 0.03 |
| North | 24,784 (9.1%) | 2,266 (10.7%) | 0.05 |
| Ottawa | 14,974 (5.5%) | 1,417 (6.7%) | 0.05 |
| Peel | 15,584 (5.7%) | 920 (4.3%) | 0.06 |
| South West | 39,021 (14.3%) | 2,891 (13.6%) | 0.02 |
| Toronto | 60,740 (22.2%) | 4,026 (18.9%) | 0.08 |
| York | 18,182 (6.6%) | 1,066 (5.0%) | 0.07 |
| Missing | 1,171 (0.4%) | 42 (0.2%) | 0.04 |
| LTC facility in outbreak at time of test | 125,449 (45.9%) | 12,731 (59.8%) | 0.28 |
| Prior positive SARS-CoV-test (>90 days) | 44,910 (16.4%) | 1,812 (8.5%) | 0.24 |
| Week of test^d^ |  |  |  |
| 30 Dec to 05 Jan | 30,196 (11.0%) | 1,959 (9.2%) | 0.06 |
| 06 Jan to 12 Jan | 30,358 (11.1%) | 2,491 (11.7%) | 0.02 |
| 13 Jan to 19 Jan | 23,131 (8.5%) | 1,944 (9.1%) | 0.02 |
| 20 Jan to 26 Jan | 19,817 (7.2%) | 1,528 (7.2%) | 0.00 |
| 27 Jan to 02 Feb | 15,651 (5.7%) | 981 (4.6%) | 0.05 |
| 03 Feb to 09 Feb | 10,406 (3.8%) | 528 (2.5%) | 0.08 |
| 10 Feb to 16 Feb | 6,951 (2.5%) | 267 (1.3%) | 0.09 |
| 17 Feb to 23 Feb | 5,859 (2.1%) | 187 (0.9%) | 0.10 |
| 24 Feb to 02 Mar | 6,052 (2.2%) | 173 (0.8%) | 0.11 |
| 03 Mar to 09 Mar | 5,207 (1.9%) | 172 (0.8%) | 0.09 |
| 10 Mar to 16 Mar | 5,480 (2.0%) | 193 (0.9%) | 0.09 |
| 17 Mar to 23 Mar | 5,618 (2.1%) | 204 (1.0%) | 0.09 |
| 24 Mar to 30 Mar | 6,479 (2.4%) | 285 (1.3%) | 0.08 |
| 31 Mar to 6 Apr | 7,841 (2.9%) | 457 (2.1%) | 0.05 |
| 7 Apr to 13 Apr | 9,439 (3.5%) | 583 (2.7%) | 0.04 |
| 14 Apr to 20 Apr | 9,035 (3.3%) | 800 (3.8%) | 0.02 |
| 21 Apr to 27 Apr | 9,320 (3.4%) | 947 (4.5%) | 0.05 |
| 28 Apr to 04 May | 7,363 (2.7%) | 797 (3.7%) | 0.06 |
| 5 May to 11 May | 5,794 (2.1%) | 640 (3.0%) | 0.06 |
| 12 May to 18 May | 6,403 (2.3%) | 574 (2.7%) | 0.02 |
| 19 May to 25 May | 4,981 (1.8%) | 346 (1.6%) | 0.01 |
| 26 May to 01 June | 4,345 (1.6%) | 343 (1.6%) | 0.00 |
| 02 June to 08 June | 3,781 (1.4%) | 230 (1.1%) | 0.03 |
| 09 June to 15 June | 3,712 (1.4%) | 176 (0.8%) | 0.05 |
| 16 June to 22 June | 3,450 (1.3%) | 233 (1.1%) | 0.02 |
| 23 June to 29 June | 3,784 (1.4%) | 258 (1.2%) | 0.02 |
| 30 June to 06 July | 4,012 (1.5%) | 360 (1.7%) | 0.02 |
| 07 July to 13 July | 4,861 (1.8%) | 669 (3.1%) | 0.09 |
| 14 July to 20 July | 5,590 (2.0%) | 1,133 (5.3%) | 0.17 |
| 21 July to 27 July | 5,322 (1.9%) | 926 (4.4%) | 0.14 |
| 28 July to 03 Aug | 3,228 (1.2%) | 891 (4.2%) | 0.19 |
| Number of comorbidities (mean, SD^d^) | 4.13 ± 1.57 | 4.10 ± 1.55 | 0.02 |
| Type of comorbidity |  |  |  |
| Immunocompromised | 25,845 (9.5%) | 1,986 (9.3%) | 0.00 |
| Chronic respiratory disease | 99,693 (36.5%) | 7,695 (36.2%) | 0.01 |
| Chronic heart disease | 103,803 (38.0%) | 7,699 (36.2%) | 0.04 |
| Hypertension | 224,114 (82.0%) | 17,407 (81.8%) | 0.00 |
| Diabetes | 110,357 (40.4%) | 8,243 (38.7%) | 0.03 |
| Autoimmune disorders | 22,674 (8.3%) | 1,662 (7.8%) | 0.02 |
| Chronic kidney disease or dialysis^e^ | 48,963 (17.9%) | 3,436 (16.2%) | 0.05 |
| Advanced liver disease | 7,435 (2.7%) | 505 (2.4%) | 0.02 |
| Dementia | 219,027 (80.1%) | 17,538 (82.4%) | 0.06 |
| History of stroke or transient  ischemic attack | 48,868 (17.9%) | 3,629 (17.1%) | 0.02 |
| Frailty | 219,317 (80.2%) | 17,381 (81.7%) | 0.04 |

^a^Proportion reported, unless stated otherwise.

^b^SD=standardized difference. Standardized differences of >0.10 are considered clinically relevant. Comparing Omicron cases to test-negative controls.

^c^Standard deviation.

^d^December 30, 31 in 2021 and remaining dates in 2022.

^e^Chronic kidney disease in the prior 5 years or dialysis for 3 consecutive months

**Table S2:** Descriptive characteristics of long-term care (LTC) residents tested for SARS-CoV-2 between December 30, 2021 and August 3, 2022 in Ontario, Canada, comparing individuals with a fourth dose to individuals with a third dose ≥84 days ago*

|  | **Received 3^rd^ dose ≥84 days prior to test, n (%)^a^** | **Received 4th dose <84 days prior to test, n (%)^a^** | **SD^b^** | **Received 4th dose 84-111 days prior to test, n(%)^a^** | **SD^b^** | **Received 4th dose 112-139 days prior to test, n (%)^a^** | **SD^b^** | **Received 4th dose 140-167 days prior to test, n (%)^a^** | **SD^b^** | **Received 4th dose ≥168 days prior to test, n (%)^a^** | **SD^b^** |
| --- | --- | --- | --- | --- | --- | --- | --- | --- | --- | --- | --- |
| **Total** | 98,483 | 81,431 |  | 23,365 |  | 14,347 |  | 11,178 |  | 12,999 |  |
| Characteristics |  |  |  |  |  |  |  |  |  |  |  |
| Age (years),  mean (standard  deviation) | 83.88 ± 9.50 | 83.53 ± 9.55 | 0.04 | 83.36 ± 9.61 | 0.05 | 83.25 ± 9.74 | 0.07 | 82.57 ± 9.87 | 0.13 | 83.22 ± 9.73 | 0.07 |
| 60 to 69 | 9,362 (9.5%) | 8,219 (10.1%) | 0.02 | 2,420 (10.4%) | 0.03 | 1,562 (10.9%) | 0.05 | 1,397 (12.5%) | 0.10 | 1,404 (10.8%) | 0.04 |
| 70 to 79 | 20,095 (20.4%) | 17,011 (20.9%) | 0.01 | 5,065 (21.7%) | 0.03 | 3,092 (21.6%) | 0.03 | 2,571 (23.0%) | 0.06 | 2,868 (22.1%) | 0.04 |
| ≥80 | 69,026 (70.1%) | 56,201 (69.0%) | 0.02 | 15,880 (68.0%) | 0.05 | 9,693 (67.6%) | 0.05 | 7,210 (64.5%) | 0.12 | 8,727 (67.1%) | 0.06 |
| Male sex | 30,873 (31.3%) | 25,645 (31.5%) | 0.00 | 7,209 (30.9%) | 0.01 | 4,496 (31.3%) | 0.00 | 3,681 (32.9%) | 0.03 | 3,861 (29.7%) | 0.04 |
| Public health  unit region |  |  |  |  |  |  |  |  |  |  |  |
| Central East | 7,807 (7.9%) | 6,299 (7.7%) | 0.01 | 1,339 (5.7%) | 0.09 | 854 (6.0%) | 0.08 | 708 (6.3%) | 0.06 | 833 (6.4%) | 0.06 |
| Central West | 18,411 (18.7%) | 13,093 (16.1%) | 0.07 | 3,484 (14.9%) | 0.10 | 1,838 (12.8%) | 0.16 | 1,683 (15.1%) | 0.10 | 1,844 (14.2%) | 0.12 |
| Durham | 3,448 (3.5%) | 3,277 (4.0%) | 0.03 | 887 (3.8%) | 0.02 | 479 (3.3%) | 0.01 | 385 (3.4%) | 0.00 | 495 (3.8%) | 0.02 |
| Eastern | 7,786 (7.9%) | 8,972 (11.0%) | 0.11 | 2,812 (12.0%) | 0.14 | 1,449 (10.1%) | 0.08 | 823 (7.4%) | 0.02 | 1,154 (8.9%) | 0.04 |
| North | 8,742 (8.9%) | 8,834 (10.8%) | 0.07 | 2,274 (9.7%) | 0.03 | 1,114 (7.8%) | 0.04 | 874 (7.8%) | 0.04 | 688 (5.3%) | 0.14 |
| Ottawa | 4,669 (4.7%) | 4,851 (6.0%) | 0.05 | 1,774 (7.6%) | 0.12 | 884 (6.2%) | 0.06 | 767 (6.9%) | 0.09 | 927 (7.1%) | 0.10 |
| Peel | 5,409 (5.5%) | 3,725 (4.6%) | 0.04 | 1,168 (5.0%) | 0.02 | 685 (4.8%) | 0.03 | 675 (6.0%) | 0.02 | 783 (6.0%) | 0.02 |
| South West | 13,305 (13.5%) | 12,281 (15.1%) | 0.04 | 3,689 (15.8%) | 0.06 | 2,283 (15.9%) | 0.07 | 1,960 (17.5%) | 0.11 | 2,125 (16.3%) | 0.08 |
| Toronto | 21,349 (21.7%) | 14,881 (18.3%) | 0.09 | 4,587 (19.6%) | 0.05 | 3,846 (26.8%) | 0.12 | 2,768 (24.8%) | 0.07 | 3,586 (27.6%) | 0.14 |
| York | 7,166 (7.3%) | 4,952 (6.1%) | 0.05 | 1,302 (5.6%) | 0.07 | 889 (6.2%) | 0.04 | 502 (4.5%) | 0.12 | 556 (4.3%) | 0.13 |
| Missing | 391 (0.4%) | 266 (0.3%) | 0.01 | 49 (0.2%) | 0.03 | 26 (0.2%) | 0.04 | 33 (0.3%) | 0.02 | 8 (0.1%) | 0.07 |
| LTC facility in  outbreak at  time of test | 54,542 (55.4%) | 33,885 (41.6%) | 0.28 | 9,735 (41.7%) | 0.28 | 4,256 (29.7%) | 0.54 | 2,374 (21.2%) | 0.75 | 4,459 (34.3%) | 0.43 |
| Prior positive  SARS-CoV-  test (>90 days) | 16,440 (16.7%) | 11,987 (14.7%) | 0.05 | 3,565 (15.3%) | 0.04 | 2,437 (17.0%) | 0.01 | 2,051 (18.3%) | 0.04 | 2,613 (20.1%) | 0.09 |
| Week of test^d^ |  |  |  |  |  |  |  |  |  |  |  |
| 30 Dec to 05 Jan | 22,436 (22.8%) | 227 (0.3%) | 0.75 | 0 (0.0%) | 0.77 | 0 (0.0%) | 0.77 | 0 (0.0%) | 0.77 | 0 (0.0%) | 0.77 |
| 06 Jan to 12 Jan | 22,080 (22.4%) | 1,603 (2.0%) | 0.66 | 0 (0.0%) | 0.76 | 0 (0.0%) | 0.76 | 0 (0.0%) | 0.76 | 0 (0.0%) | 0.76 |
| 13 Jan to 19 Jan | 14,048 (14.3%) | 4,151 (5.1%) | 0.31 | 0 (0.0%) | 0.58 | 0 (0.0%) | 0.58 | 0 (0.0%) | 0.58 | 0 (0.0%) | 0.58 |
| 20 Jan to 26 Jan | 9,505 (9.7%) | 6,396 (7.9%) | 0.06 | 0 (0.0%) | 0.46 | 0 (0.0%) | 0.46 | 0 (0.0%) | 0.46 | 0 (0.0%) | 0.46 |
| 27 Jan to 02 Feb | 5,167 (5.2%) | 7,384 (9.1%) | 0.15 | 0 (0.0%) | 0.33 | 0 (0.0%) | 0.33 | 0 (0.0%) | 0.33 | 0 (0.0%) | 0.33 |
| 03 Feb to 09 Feb | 2,569 (2.6%) | 5,628 (6.9%) | 0.20 | 0 (0.0%) | 0.23 | 0 (0.0%) | 0.23 | 0 (0.0%) | 0.23 | 0 (0.0%) | 0.23 |
| 10 Feb to 16 Feb | 1,235 (1.3%) | 4,345 (5.3%) | 0.23 | 0 (0.0%) | 0.16 | 0 (0.0%) | 0.16 | 0 (0.0%) | 0.16 | 0 (0.0%) | 0.16 |
| 17 Feb to 23 Feb | 900 (0.9%) | 3,845 (4.7%) | 0.23 | 0 (0.0%) | 0.14 | 0 (0.0%) | 0.14 | 0 (0.0%) | 0.14 | 0 (0.0%) | 0.14 |
| 24 Feb to 02Mar | 977 (1.0%) | 3,927 (4.8%) | 0.23 | 0 (0.0%) | 0.14 | 0 (0.0%) | 0.14 | 0 (0.0%) | 0.14 | 0 (0.0%) | 0.14 |
| 03 Mar to 09 Mar | 766 (0.8%) | 3,541 (4.3%) | 0.23 | 0 (0.0%) | 0.13 | 0 (0.0%) | 0.13 | 0 (0.0%) | 0.13 | 0 (0.0%) | 0.13 |
| 10 Mar to 16 Mar | 844 (0.9%) | 3,920 (4.8%) | 0.24 | 0 (0.0%) | 0.13 | 0 (0.0%) | 0.13 | 0 (0.0%) | 0.13 | 0 (0.0%) | 0.13 |
| 17 Mar to 23 Mar | 900 (0.9%) | 4,187 (5.1%) | 0.25 | 0 (0.0%) | 0.14 | 0 (0.0%) | 0.14 | 0 (0.0%) | 0.14 | 0 (0.0%) | 0.14 |
| 24 Mar to 30 Mar | 1,105 (1.1%) | 4,903 (6.0%) | 0.27 | 35 (0.1%) | 0.12 | 0 (0.0%) | 0.15 | 0 (0.0%) | 0.15 | 0 (0.0%) | 0.15 |
| 31 Mar to 6 Apr | 1,438 (1.5%) | 5,587 (6.9%) | 0.27 | 486 (2.1%) | 0.05 | 0 (0.0%) | 0.17 | 0 (0.0%) | 0.17 | 0 (0.0%) | 0.17 |
| 7 Apr to 13 Apr | 1,547 (1.6%) | 5,636 (6.9%) | 0.27 | 2,045 (8.8%) | 0.33 | 0 (0.0%) | 0.18 | 0 (0.0%) | 0.18 | 0 (0.0%) | 0.18 |
| 14 Apr to 20 Apr | 1,651 (1.7%) | 3,921 (4.8%) | 0.18 | 3,558 (15.2%) | 0.50 | 0 (0.0%) | 0.18 | 0 (0.0%) | 0.18 | 0 (0.0%) | 0.18 |
| 21 Apr to 27 Apr | 1,662 (1.7%) | 2,882 (3.5%) | 0.12 | 4,890 (20.9%) | 0.64 | 46 (0.3%) | 0.14 | 0 (0.0%) | 0.19 | 0 (0.0%) | 0.19 |
| 28 Apr to 04 May | 1,321 (1.3%) | 1,772 (2.2%) | 0.06 | 4,063 (17.4%) | 0.57 | 449 (3.1%) | 0.12 | 0 (0.0%) | 0.16 | 0 (0.0%) | 0.16 |
| 5 May to 11 May | 1,027 (1.0%) | 1,158 (1.4%) | 0.03 | 2,559 (11.0%) | 0.43 | 1,240 (8.6%) | 0.36 | 0 (0.0%) | 0.15 | 0 (0.0%) | 0.15 |
| 12 May to 18 May | 1,012 (1.0%) | 1,256 (1.5%) | 0.05 | 1,928 (8.3%) | 0.35 | 2,371 (16.5%) | 0.57 | 0 (0.0%) | 0.14 | 0 (0.0%) | 0.14 |
| 19 May to 25 May | 807 (0.8%) | 792 (1.0%) | 0.02 | 822 (3.5%) | 0.19 | 2,531 (17.6%) | 0.61 | 43 (0.4%) | 0.06 | 0 (0.0%) | 0.13 |
| 26 May to 01 June | 684 (0.7%) | 745 (0.9%) | 0.02 | 544 (2.3%) | 0.13 | 2,182 (15.2%) | 0.56 | 261 (2.3%) | 0.13 | 0 (0.0%) | 0.12 |
| 02 June to 08 June | 536 (0.5%) | 518 (0.6%) | 0.01 | 329 (1.4%) | 0.09 | 1,646 (11.5%) | 0.47 | 730 (6.5%) | 0.33 | 0 (0.0%) | 0.10 |
| 09 June to 15 June | 514 (0.5%) | 449 (0.6%) | 0.00 | 303 (1.3%) | 0.08 | 1,043 (7.3%) | 0.35 | 1,359 (12.2%) | 0.49 | 0 (0.0%) | 0.10 |
| 16 June to 22 June | 408 (0.4%) | 418 (0.5%) | 0.01 | 253 (1.1%) | 0.08 | 567 (4.0%) | 0.24 | 1,815 (16.2%) | 0.60 | 17 (0.1%) | 0.05 |
| 23 June to 29 June | 494 (0.5%) | 401 (0.5%) | 0.00 | 270 (1.2%) | 0.07 | 506 (3.5%) | 0.22 | 1,935 (17.3%) | 0.62 | 225 (1.7%) | 0.12 |
| 30 June to 06 July | 496 (0.5%) | 378 (0.5%) | 0.01 | 238 (1.0%) | 0.06 | 323 (2.3%) | 0.15 | 1,628 (14.6%) | 0.55 | 1,062 (8.2%) | 0.38 |
| 07 July to 13 July | 535 (0.5%) | 445 (0.5%) | 0.00 | 275 (1.2%) | 0.07 | 367 (2.6%) | 0.16 | 1,292 (11.6%) | 0.47 | 2,321 (17.9%) | 0.63 |
| 14 July to 20 July | 721 (0.7%) | 417 (0.5%) | 0.03 | 301 (1.3%) | 0.06 | 439 (3.1%) | 0.17 | 940 (8.4%) | 0.37 | 3,538 (27.2%) | 0.83 |
| 21 July to 27 July | 639 (0.6%) | 369 (0.5%) | 0.03 | 279 (1.2%) | 0.06 | 387 (2.7%) | 0.16 | 828 (7.4%) | 0.35 | 3,409 (26.2%) | 0.81 |
| 28 July to 03 Aug | 459 (0.5%) | 230 (0.3%) | 0.03 | 187 (0.8%) | 0.04 | 250 (1.7%) | 0.12 | 347 (3.1%) | 0.20 | 2,427 (18.7%) | 0.65 |
| Number of  comorbidities  (mean, SD)^c^ | 4.10 ± 1.56 | 4.10 ± 1.57 | 0.00 | 4.06 ± 1.57 | 0.02 | 4.08 ± 1.59 | 0.01 | 4.08 ± 1.60 | 0.01 | 3.99 ± 1.58 | 0.07 |
| Type of  comorbidity |  |  |  |  |  |  |  |  |  |  |  |
| Immuno-  compromised | 8,765 (8.9%) | 7,397 (9.1%) | 0.01 | 2,123 (9.1%) | 0.01 | 1,342 (9.4%) | 0.02 | 1,067 (9.5%) | 0.02 | 1,090 (8.4%) | 0.02 |
| Chronic  Respiratory  disease | 35,213 (35.8%) | 30,075 (36.9%) | 0.02 | 8,599 (36.8%) | 0.02 | 5,200 (36.2%) | 0.01 | 4,078 (36.5%) | 0.02 | 4,655 (35.8%) | 0.00 |
| Chronic heart  disease | 36,536 (37.1%) | 30,104 (37.0%) | 0.00 | 8,558 (36.6%) | 0.01 | 5,410 (37.7%) | 0.01 | 4,174 (37.3%) | 0.01 | 4,491 (34.5%) | 0.05 |
| Hypertension | 80,888 (82.1%) | 66,400 (81.5%) | 0.02 | 18,979 (81.2%) | 0.02 | 11,650 (81.2%) | 0.02 | 9,140 (81.8%) | 0.01 | 10,447 (80.4%) | 0.05 |
| Diabetes | 39,036 (39.6%) | 32,371 (39.8%) | 0.00 | 9,344 (40.0%) | 0.01 | 5,822 (40.6%) | 0.02 | 4,631 (41.4%) | 0.04 | 5,197 (40.0%) | 0.01 |
| Autoimmune  disorders | 8,143 (8.3%) | 6,883 (8.5%) | 0.01 | 1,878 (8.0%) | 0.01 | 1,189 (8.3%) | 0.00 | 985 (8.8%) | 0.02 | 1,070 (8.2%) | 0.00 |
| Chronic kidney  disease^e^ | 16,463 (16.7%) | 13,901 (17.1%) | 0.01 | 3,909 (16.7%) | 0.00 | 2,481 (17.3%) | 0.02 | 1,995 (17.8%) | 0.03 | 2,053 (15.8%) | 0.03 |
| Advanced liver  disease | 2,490 (2.5%) | 2,114 (2.6%) | 0.00 | 610 (2.6%) | 0.01 | 396 (2.8%) | 0.01 | 339 (3.0%) | 0.03 | 389 (3.0%) | 0.03 |
| Dementia | 80,664 (81.9%) | 65,473 (80.4%) | 0.04 | 18,628 (79.7%) | 0.06 | 11,241 (78.4%) | 0.09 | 8,675 (77.6%) | 0.11 | 10,389 (79.9%) | 0.05 |
| History of stroke  or transient  ischemic attack | 17,415 (17.7%) | 14,427 (17.7%) | 0.00 | 4,171 (17.9%) | 0.00 | 2,673 (18.6%) | 0.02 | 2,058 (18.4%) | 0.02 | 2,334 (18.0%) | 0.01 |
| Frailty | 78,131 (79.3%) | 64,404 (79.1%) | 0.01 | 18,177 (77.8%) | 0.04 | 11,116 (77.5%) | 0.05 | 8,422 (75.3%) | 0.10 | 9,814 (75.5%) | 0.09 |

^*^Note, not unique by person; individuals with multiple negative tests were included more than once.

^a^Proportion reported, unless stated otherwise.

^b^SD=standardized difference. Standardized differences of >0.10 are considered clinically relevant. Comparing vaccinated individuals across different doses with unvaccinated individuals.

^c^Standard deviation

^d^Dec 30, 31 in 2021, all other dates in 2022.

^e^Chronic kidney disease in the prior 5 years or dialysis for 3 consecutive months.

**Table S3:** Marginal effectiveness of a fourth dose of mRNA COVID-19 vaccine against Omicron outcomes among long-term care residents in Ontario, Canada, compared to residents who received a third dose ≥84 days ago

| **Outcome** | **SARS-CoV-2-negative controls, n** | **Omicron-positive cases, n** | **Marginal effectiveness, relative to third dose**  **≥84 days ago**  **% (95% CI)** |
| --- | --- | --- | --- |
| Infection |  |  |  |
| <84 days | 77,695 | 3,736 | 23 (17, 29) |
| 84-111 days | 21,450 | 1,915 | 13 (3, 21) |
| 112-139 days | 13,253 | 1,094 | 7 (-6, 18) |
| 140-167 days | 10,223 | 955 | 3 (-12, 16) |
| ≥168 days | 10,855 | 2,144 | -24 (-41, -9) |
| Symptomatic infection |  |  |  |
| <84 days | 3,088 | 698 | 36 (26, 44) |
| 84-111 days | 939 | 438 | 18 (1, 32) |
| 112-139 days | 634 | 270 | -5 (-35, 18) |
| 140-167 days | 563 | 211 | 10 (-15, 30) |
| ≥168 days | 485 | 477 | -15 (-49, 11) |
| Severe outcomes |  |  |  |
| <84 days | 3,088 | 164 | 37 (24, 48) |
| 84-111 days | 939 | 78 | 24 (3, 41) |
| 112-139 days | 634 | 46 | 28 (-4, 50) |
| 140-167 days | 563 | 52 | 23 (-7, 45) |
| ≥168 days | 485 | 104 | 6 (-29, 32) |

CI = confidence interval.

**Table S4:** Vaccine effectiveness of 2, 3, and 4 doses of an mRNA COVID-19 vaccine against Omicron outcomes among long-term care residents in Ontario, Canada, compared to unvaccinated residents

| **Outcome** | **SARS-CoV-2-negative controls, n** | **Omicron-positive cases, n** | **Vaccine effectiveness (95% CI)** |
| --- | --- | --- | --- |
| Infection |  |  |  |
| 2 doses | 12,472 | 1,442 | 3 (-7, 12) |
| 3 doses (<84 days) | 28,133 | 1,842 | 38 (32, 43) |
| 3 doses (≥84 days) | 91,251 | 7,232 | 34 (28, 39) |
| 4 doses (<84 days) | 77,695 | 3,736 | 49 (44, 54) |
| 4 doses (84-111 days) | 21,450 | 1,915 | 42 (36, 48) |
| 4 doses (112-139 days) | 13,253 | 1,094 | 38 (29, 46) |
| 4 doses (140-167 days) | 10,223 | 955 | 36 (26, 44) |
| 4 doses (≥168 days) | 10,855 | 2,144 | 18 (5, 28) |
| Symptomatic infection |  |  |  |
| 2 doses | 367 | 292 | 21 (1, 38) |
| 3 doses (<84 days) | 774 | 273 | 59 (48, 68) |
| 3 doses (≥84 days) | 2,772 | 1,321 | 51 (42, 60) |
| 4 doses (<84 days) | 3,088 | 698 | 69 (61, 75) |
| 4 doses (84-111 days) | 939 | 438 | 60 (49, 69) |
| 4 doses (112-139 days) | 634 | 270 | 49 (32, 62) |
| 4 doses (140-167 days) | 563 | 211 | 56 (41, 67) |
| 4 doses (≥168 days) | 485 | 477 | 44 (24, 59) |
| Severe outcomes |  |  |  |
| 2 doses | 367 | 126 | 43 (24, 58) |
| 3 doses (<84 days) | 774 | 97 | 77 (68, 83) |
| 3 doses (≥84 days) | 2,772 | 418 | 72 (64, 78) |
| 4 doses (<84 days) | 3,088 | 164 | 82 (77, 86) |
| 4 doses (84-111 days) | 939 | 78 | 79 (71, 84) |
| 4 doses (112-139 days) | 634 | 46 | 80 (70, 86) |
| 4 doses (140-167 days) | 563 | 52 | 78 (69, 85) |
| 4 doses (≥168 days) | 485 | 104 | 74 (62, 82) |

CI = confidence interval.
